# Thalamic stimulation induced changes in network connectivity and excitability in epilepsy

**DOI:** 10.1101/2024.03.03.24303480

**Authors:** Nicholas M. Gregg, Gabriela Ojeda Valencia, Tereza Pridalova, Harvey Huang, Vaclav Kremen, Brian N. Lundstrom, Jamie J. Van Gompel, Kai J. Miller, Gregory A. Worrell, Dora Hermes

**Author notes:** Correspondence to: Nicholas Gregg, MD 200 First Street South Wester, Rochester, MN, 55905. Dora Hermes, PhD Gregory Worrell, MD PhD. contributed equally (Gregory A. Worrell, Dora Hermes).

## Abstract

**Objective:** The effects of deep brain stimulation (DBS) manifest across multiple timescales, spanning seconds to months, and involve direct electrical effects, neuroplasticity, and network reorganization. In epilepsy, the delayed impact of DBS on seizures presents challenges for optimization. Single-pulse stimulation and resulting brain stimulation evoked potentials (BSEPs) provide a means to assess effective connectivity and network excitability. This study integrates BSEPs and short trials of DBS during stereoelectroencephalography (sEEG) to map seizure network engagement, modulate network dynamics, and monitor excitability and interictal abnormalities, for biomarker informed neuromodulation.

**Methods:** Ten individuals with drug resistant epilepsy undergoing clinical sEEG were enrolled in this retrospective cohort study of epilepsy neuromodulation biomarkers. Each patient underwent a trial of high frequency (145 Hz) thalamic DBS. BSEPs were acquired before and after DBS trials. Baseline BSEP amplitude assessed seizure network engagement, and modulation of amplitude (pre vs. post DBS) assessed change in network excitability. Interictal epileptiform discharges were tracked by an automated classifier.

**Results:** Baseline BSEPs delineated distinct patterns of network engagement between thalamic subfields, with maximal frontotemporal engagement achieved with stimulation of the anterior nucleus of the thalamus-ventral anterior nucleus junction. DBS delivered for >1.5 hours reduced BSEP amplitudes compared to baseline, and the degree of modulation correlated with baseline connectivity strength. Shorter DBS trials did not induce reliable BSEP amplitude suppression, but did immediately suppress interictal epileptiform discharge rates in well-connected seizure networks.

**Interpretation:** BSEPs and trials of DBS during sEEG provide novel network biomarkers to evaluate the modulation of large-scale networks across multiple timescales, advancing biomarker informed neuromodulation.

## Introduction

Deep brain stimulation (DBS) is a viable treatment for a variety of drug-resistant neurological conditions. However, the mechanisms by which DBS modulates large-scale brain networks remain unresolved. Clinical effects of DBS manifest over different timescales, with variability in response rates and symptom improvement across disorders. Particularly favorable outcomes are observed in disorders with well-defined pathological circuits and when DBS effects on clinical symptoms are immediate, allowing for rapid screening and parameter optimization (e.g. Parkinson’s disease tremor and essential tremor)^1^.

In contrast, in conditions such as epilepsy^2^, central pain^3^, dystonia^4^, and neuropsychiatric conditions^5^, the DBS related effects require neuroplasticity or reorganization, which can take hours to months to observe^6^, with generally less favorable outcomes when compared to tremor disorders. Epilepsy, a brain network disorder^7–9^, is notable for cross-subject heterogeneity with subject-specific seizure network (SN) structures^10^. Furthermore, the primary clinical manifestation of epilepsy^11^—epileptic seizures—have highly dynamic risk profiles^12^ with inter-seizure intervals ranging from hours to months. These features present challenges for targeting DBS electrodes to engage and individual’s SN and tuning stimulation parameters across a massive potential stimulation parameter space.

In this study, we present an electrophysiological biomarker that rapidly quantifies network-level changes in connectivity and excitability, with the goal of guiding therapeutic neuromodulation for epilepsy. During stereotactic electroencephalography (sEEG) for individuals with drug resistant focal epilepsy, a number of multi-contact penetrating depth electrodes are used to map and characterize pathological networks and identify eloquent structures. Single pulse electrical stimulation can evoke characteristic responses in connected regions, here termed brain stimulation evoked potentials (BSEPs), which map effective connectivity throughout the network^13,14^. Effective connectivity reflects the causal influence of a stimulated site on other brain regions, and analysis of BSEPs over time can be used to quantify changes in network excitability^15–17^.

Interictal epileptiform discharges (IEDs), a hallmark subclinical electrophysiological feature of epilepsy^11^, are brief paroxysmal bursts of hypersynchronous neuronal firing, detectable via EEG. In this study, we test whether BSEPs—acting as biomarkers of network effective connectivity and excitability—can quantify meaningful neural changes induced by high frequency (HF) thalamic DBS, a proven therapy for reducing seizures in drug resistant focal epilepsy^2^. We also assess the ability of BSEPs to predict DBS modulation of SN IEDs.

## Materials and Methods

This is a retrospective series evaluating the effects of clinical HF-DBS on electrophysiological biomarkers during sEEG monitoring performed as part of the evaluation for drug resistant focal epilepsy. Electrical stimulation for mapping thalamocortical networks before and after HF-DBS was performed under Mayo Clinic Institutional Review Board approved protocol (IRB #15- 006530). Patients were enrolled at the Mayo Clinic between June 2020 and December 2022. All patients in the study completed a written informed consent. Thalamic leads were placed per clinical care and were not influenced by this study. The thalamic lead targets were selected by the multi-disciplinary clinical surgical epilepsy conference consensus recommendations. Thalamic targets were patient specific and guided by the hypothesized SN. Clinical stimulation trials are completed to assess stimulation effects on the SN and tolerability, with the advantage of high-quality local field potential recordings from distributed brain regions (up to 256 recording contacts, Natus Medical Inc. Quantum amplifier).

Patient characteristics are listed in Table 1. There was some heterogeneity in the clinical HF-DBS parameters. Patient 1 completed bilateral anterior thalamus HF-DBS trial stimulation. Patient 6 underwent single pulse stimulation at baseline, after 1 hour of HF-DBS, and again after 5.8 hours of HF-DBS (listed HF-DBS durations reflect only the active phase of duty-cycle stimulation, e.g. Patient 4 with a 1 min. on 3 min. off duty-cycle received 4.3 hr. of active stimulation, interleaved with 12.9 hr. of off-phase time).

**Table 1.** Patient characteristics. The duration of thalamic high frequency deep brain stimulation (HF-DBS) is the active stimulation time (does not include off-phase of duty-cycle stimulation). Patient 6 completed single pulse stimulation after 1-hour and 5.8-hours of HF-DBS. L=left. R=right. FCD=focal cortical dysplasia. ANT=anterior nucleus of the thalamus. Thalamic nuclei abbreviations, Krauth/Morel atlas^23^. AM=anteromedial nucleus. AV=anteroventral nucleus. VA=ventral anterior nucleus. VL=ventral lateral nucleus, MTT=mammillothalamic tract, PuM=medial pulvinar. MD=mediodorsal nucleus. Duty-cycle (on-period and off-period) noted in minutes. NC=no change in medication regimen during the single pulse and HF-DBS period. LEV=levetiracetam. LTG=lamotrigine. ZNS=zonisamide. *Additional cortical stimulation site (Supplemental Table 2)

| Patient | Age | Sex | Seizure onset | Thalamic stim. location | Stimulation parameters | Duration thalamic HF-DBS | Imaging | Med. change |
| --- | --- | --- | --- | --- | --- | --- | --- | --- |
| 1 | 17 | m | Diffuse bifrontal, left hemisphere lead in | R-AM, AV, VApc; L-AM, AV, VApc | 145 Hz, 90 μs, 5.7mA (total), 1m. on 1m. off | 7 hr. | Nonlesional | NC |
| 2 | 48 | m | Left middle temporal gyrus | L-AV, VApc, VLpd | 145 Hz, 90 μs, 3.6mA, continuous | 1.2 hr. | Left temporal encephalomalacia secondary to meningioma, status post resection | NC |
| 3 | 26 | m | Right hippocampus. Frequent bilateral mesial temporal IEDs. | R-AM, AV, VApc, VAmc, VLpv, MTT | 145 Hz, 200 μs, 3.7mA, 1m. on 1m. off | 1.8 hr. | Left mesial temporal atrophy. Left hemisphere PET hypometabolism. | NC |
| 4 | 12 | m | Right occipital, peri-lesion (perinatal stroke) | R-PulM | 145 Hz, 200 μs, 5.8mA, 1m. on 3m. off | 4.3 hr. | Posterior midline ulegyria, right greater than left. | change (increase LTG, restart ZNS) |
| 5 | 19 | f | Left hippocampus, peri-lesion (neoplasm) | L-VLpd | 145 Hz, 200 μs, 4.6mA, 1m. on 3m. off | 4.3 hr. | Left mesial temporal encephalomalacia from ganglioglioma, status-post resection | NC |
| 6 | 68 | f | Left neocortical and mesial temporal | L-VApc | 145 Hz, 200 μs, 4.3mA, 1m. on 3m. off | 1 hr.; 5.8 hr. | Mild global volume loss; small zone of encephalomalacia right basal ganglia | NC |
| 7 | 42 | m | Bilateral anterior cingulate | R-VApc | 145 Hz, 200 μs, 2.5mA, 1m. on 3m. off | 0.5 hr. | Multifocal, bilateral, small cortical T2/FLAIR hyperintense regions, consistent with traumatic injury | NC |
| 8 | 16 | f | Right neocortical temporal, superior frontal, and precentral gyri | R-VApc, VLpv | 145 Hz, 200 μs, 3.3mA, 1m. on 3m. off | 9.8 hr. | Nonlesional | NC |
| 9 | 7 | f | Left pre- and post-central gyri, peri-lesion (FCD) | L-VLpv | 145 Hz, 200 μs, 2.8mA, 1m. on 5m. off | 0.5 hr. | Left precentral gyrus focal cortical dysplasia | NC |
| 10 | 17 | f | Multifocal, right anterior/middle insula, amygdala, temporal neocortex | R-CL, AV, MDpc | 145 Hz, 90 μs, 3.2mA, 1m. on 5m. off | 0.8 hr. | Right frontal encephalomalacia from low-grade glioma, status post resection | Rescue IV lorazepam & LEV pre baseline |

Single pulse electrical stimulation occurred in the awake state prior to and following (within 1-hour) HF-DBS. Bipolar single pulse stimulation was delivered through neighboring contacts on the same lead, using symmetric charge balanced biphasic stimulation pulses with leading cathodal phase, delivered at 0.2 Hz, for 10-15 repetitions. Single pulse stimulation was delivered by a Natus Nicolet stimulator (Patients 1, 2, 7, 9), g.tec g.ESTIM PRO with g.HiAmp amplifier (Patients 4, 8), or Medtronic external neurostimulator 37022 (Patients 3, 5, 6, 10).

Current clamped systems (Natus and g.tec) stimuli used amplitudes of 4-6 mA, and voltage clamped (Medtronic) systems used 6-8 V (typical impedance 1-2.5 kilohms; consistent voltage used at baseline and post-DBS). The g.tec stimulator used pulse widths of 100 microseconds, while the two other systems used 200 microseconds. The Medtronic external neurostimulator was used to deliver HF (145 Hz) thalamic stimulation for all subjects.

Thalamocortical evoked potential data were first cleaned of stimuli with excessive artifact. High pass (1 Hz), low pass (170 Hz) and bandstop (for 60 Hz line noise and harmonics (120 Hz and 180 Hz)) filtering was performed with fourth order Butterworth filters, with forward-reverse filtering to correct for phase distortion. Voltage trace baseline correction consisted of subtracting the median value for window 200 to 50 milliseconds pre-stimulus from the tracing. Lastly, adjusted common average referencing was performed as previously described^18^.

To quantify the strength of effective connectivity, baseline BSEP root-mean-square (RMS) amplitude was calculated over time window 20 through 300 milliseconds post single pulse stimulus. The time window was chosen to encompass typical N1 and N2 latencies while omitting potential stimulation artifact^13^. The statistical significance of evoked potentials was assessed by paired-sample t-test, comparing RMS amplitude over this window to a pre-stimulation period of equal duration (480 to 200 milliseconds pre-stimulus, which avoids the baseline correction window). Changes in network excitability are thought to modulate the amplitude of stimulation evoked measures of effective connectivity. Here, modulation of excitability by HF stimulation was evaluated using Cohen’s *d,* as has been used previously^15^, with Cohen’s *d* effect size equal to the difference in mean evoked potential RMS amplitude (post-HF DBS vs. baseline) divided by the pooled standard deviation. A linear regression model, using Ordinary Least Squares evaluated the association between baseline effective connectivity, and HF stimulation induced changes in network excitability. The impact of short versus long durations of HF-DBS on effective connectivity was assessed by comparing the slopes of the linear regression model between the two categories, using the two-sample t-test.

SN IEDs were quantified using a validated classifier^19,20^. The SN was defined by the clinical record and iEEG review. IEDs were assessed in the awake state to avoid confounding by well-known wake/sleep-state dependent changes in IED rates. Behavioral state was determined as previously described^21^. The average spike rate of SN contacts, in the awake state, was assessed over 1 to 12 hours at baseline and 2-15 hours with active DBS. Of note, Patient 7 completed a trial of rapid-cycling HF-DBS (2 hours 7 minutes), with duty-cycle of 10 sec. on 10 sec. off, and IED rate data was drawn from this trial. This was used due to the observation by the clinical team that this patient had return of IEDs following a brief period of suppression at the 1 min. active-phase onset for each typical duty-cycle completed earlier. (Rapid cycling stimulation trial occurred after the post HF-DBS BSEPs were measured, and BSEP results were from the conventional duty-cycle regime (Table 1)).

Post-operative CT, and pre-operative T1-weighted MRI (MPRAGE) images were used for lead localization. Open source Lead DBS imaging package (v2.5.3)^22^ was used for thalamus electrode localization relative to the Krauth/Morel atlas^23^, adjusted for use with the Montreal Neurological Institute (MNI) 2009b asymmetric template space used in the Lead-DBS package. The geometric distance between stimulated electrode contacts to Krauth/Morel thalamic atlas^23^ structures was computed, and structures within 1 mm of stimulated contacts were considered to be within the volume of activated tissue^22^. For connectivity outcomes, patients were grouped into limbic and motor thalamus, motor thalamus, or medial pulvinar engagement categories.

Extra-thalamic lead localization and patient specific image rendering was performed using FreeSurfer 7^24^ and custom scripts (available at Multimodal Neuroimaging Lab (MNL) GitHub: https://github.com/MultimodalNeuroimagingLab). Contacts were localized relative to the Destrieux atlas^25^ cortical parcellation and a subcortical segmentation method (FreeSurfer Automatic Subcortical Segmentation) and clustered into orbitofrontal, cingulate, insula, neocortical temporal, mesial temporal, prefrontal, parietal, occipital, precentral, and postcentral groups^26^. Expected thalamic nucleus-cortical connectivity was assigned based on preexisting anatomical tracing and imaging work^26^ and thalamocortical BSEP amplitudes were compared between contacts within and outside of expected neuroanatomical networks; expected thalamocortical network connectivity was assigned as: AV: orbitofrontal, cingulate, insula.

AM/MTT: orbitofrontal, cingulate, insula, temporal. VA/VL: precentral and prefrontal. PuM: parietal, occipital, neocortical temporal. MD: orbitofrontal, prefrontal, temporal. Group electrode renderings in Montreal Neurological Institute (MNI) space were completed using SPM12 and custom scripts (MNL GitHub). For category-level connectivity mapping, BSEP RMS Cohen’s d values were normalized to each patient’s 90^th^ percentile value, and nonsignificant channels were zeroed. Contact positions were flipped into the left-hemisphere, as needed, and weights were projected onto the MNI cortical mesh, and a 3D Gaussian smoothing kernel (half-max radius = 7mm) was applied to create thalamic category-level (limbic and motor thalamus; motor thalamus; medial pulvinar) intensity maps (Figure 3). Analysis of interictal epileptiform discharges was performed using Python (v3.10.13, Python Software Foundation); all other analyses were performed using MATLAB (v2023a, MathWorks). Data and code to reproduce the main findings of this study are available on GitHub (https://github.com/nmgregg/Thalamic-stim-connect).

## Results

Eleven subjects with drug resistant epilepsy underwent clinical sEEG monitoring including a thalamic lead, completed a trial of thalamic HF-DBS delivered through sEEG electrodes, and had BSEPs collected prior to and following HF-DBS. One subject was excluded due to high impedance thalamic contacts and poor energy delivery during stimulation. Patient characteristics, sEEG lead location, and treatment stimulation parameters were selected based on the clinical evaluation (Table 1). The baseline and post HF-DBS BSEP amplitudes are shown in Fig. 1D for Patient 3, illustrating a reduction in evoked response amplitude after a period of HF-DBS. The change in BSEP amplitude has been argued to indicate changes in network excitability^15^.

**Figure 1.**
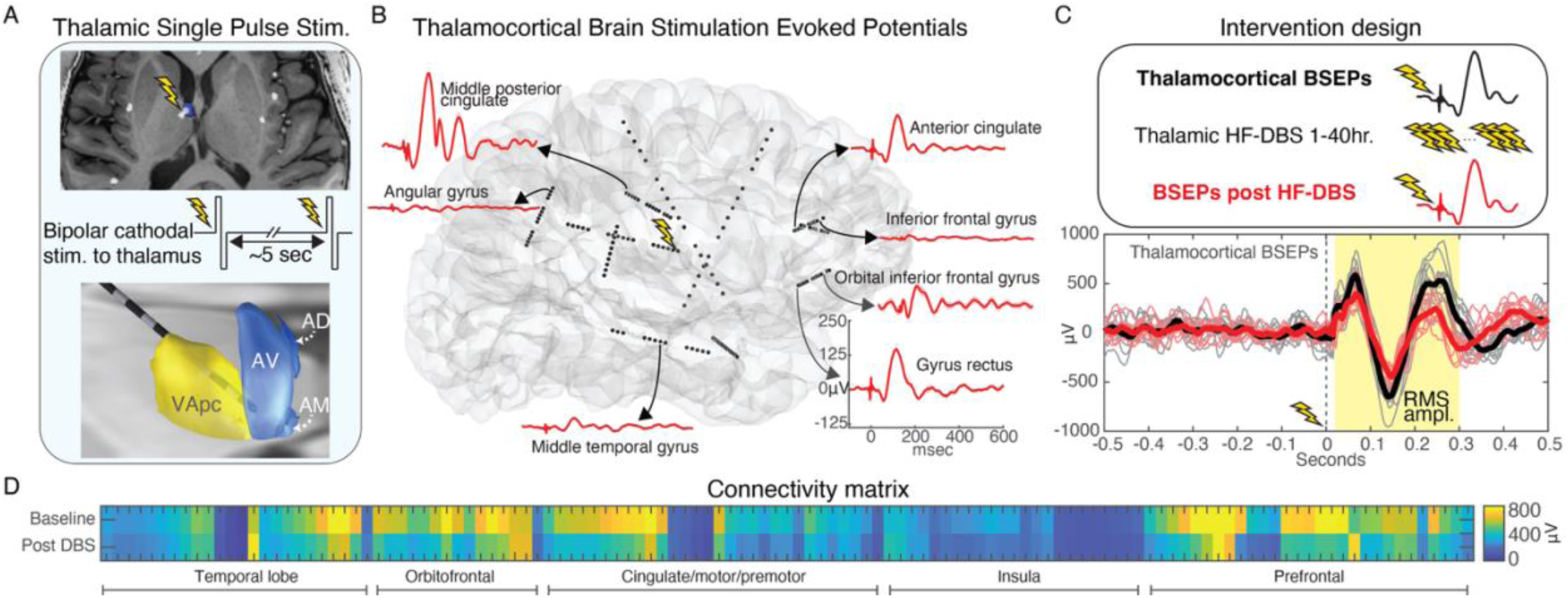
Thalamic high frequency deep brain stimulation and network effective connectivity. A) Bipolar single pulses of electrical stimulation (charge balanced symmetric square wave, leading cathodal phase) are delivered to neighboring thalamic electrode contacts. B) Single pulses delivered to the anterior nucleus of thalamus, shown in Fig. 1A (neighboring contact bipolar stimulation, contact 1 is cathode) produce characteristic evoked responses in connected brain regions, evident in sEEG electrode voltage traces (figure shows representative evoked responses; average trace from n=10 single pulse stimuli). C) Brain stimulation evoked potential (BSEP) reflecting the thalamic stimulation evoked cortical voltage response were measured at baseline and following HF-DBS. Single trial and average voltage traces are shown for baseline (black) and post-high frequency stimulation (red). The root-mean-square (RMS) amplitude of BSEPs was calculated over a window [20, 300] ms after the single pulse stimulus, for each recording contact. D) Thalamocortical effective connectivity matrix shows BSEP RMS amplitude at baseline, and post HF-DBS stimulation, across all contacts, with clear suppression of thalamocortical BSEP amplitude (excitability) over multiple regions. Data from patient 3. AD = anterodorsal nucleus. AV = anteroventral nucleus. AM = anteromedial nucleus. VApc = ventral anterior nucleus, parvocellular division. BSEP = brain stimulation evoked potential. HF-DBS = high frequency deep brain stimulation.

Thalamocortical effective connectivity was assessed in 10 patients, with a total of 11 thalamus leads (Figure 2A). Per clinical decisions, thalamus electrodes primarily targeted the anterior thalamus (n=9 subjects), with active contacts in the anterior complex (anteromedial (AM) and anteroventral (AV) nuclei), ventral group (ventral anterior (VA) and ventrolateral (VL) nuclei), and medial group (mediodorsal nucleus (MD)). One patient had electrodes in the medial pulvinar (PuM). Figure 2 shows the 11 thalamic leads in MNI template space, overlayed on the Krauth/Morel thalamus atlas^23^ (Lead-DBS imaging package (v2.5.3)^22^; BigBrain 3D human brain^27^).

**Figure 2.**
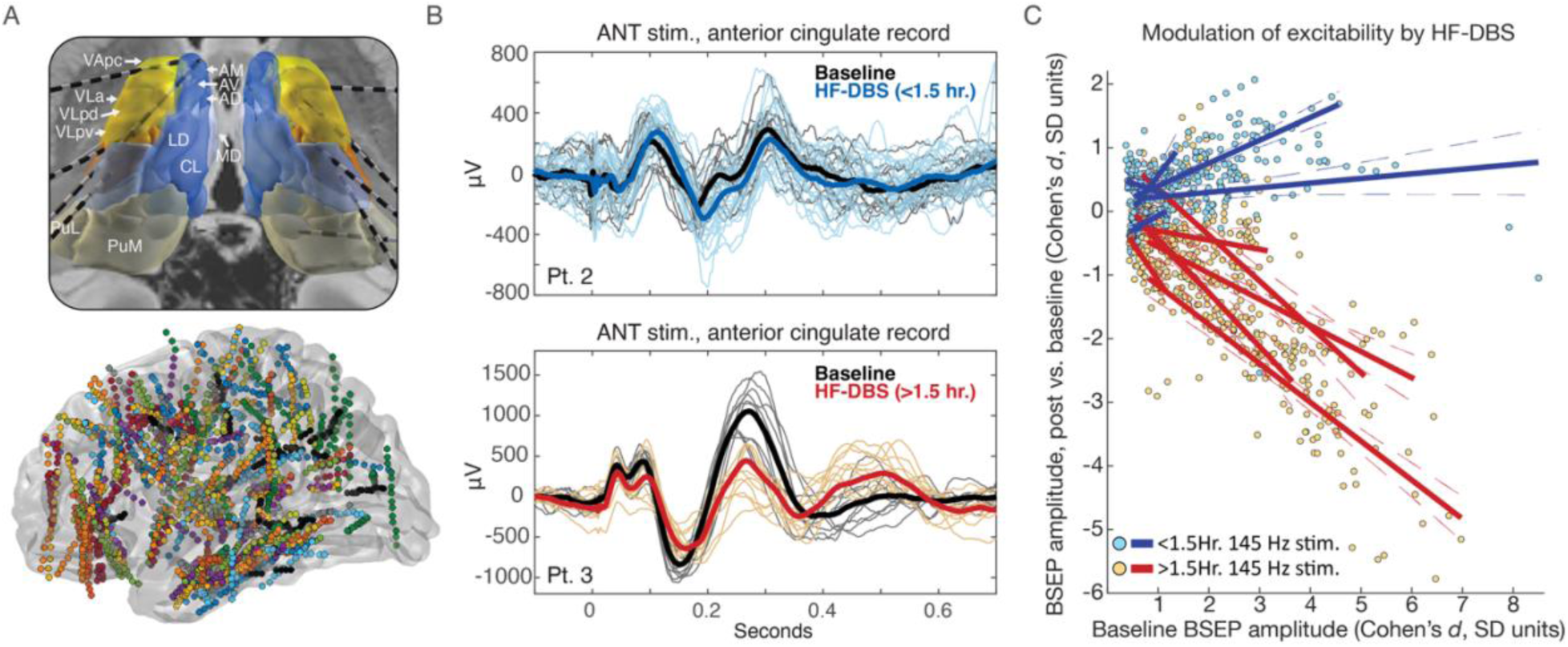
Thalamic high frequency stimulation modulates network effective connectivity and is dependent on baseline connectivity strength, and duration. A) Ten patient cohort with thalamus electrodes (top panel, Krauth/Morel thalamus atlas23) and all recording electrodes (bottom panel) shown in MNI template space (right hemisphere electrodes mirrored into left hemisphere). B) Example single trial and average thalamocortical evoked potential (EP) traces from two subjects; top panel shows BSEPs at baseline and following short duration HF-DBS, and bottom panel shows BSEPs at baseline and following long duration thalamic HF-DBS. C) Modulation of BSEP amplitude is dependent on high frequency stimulation duration(>1.5 vs. <1.5 hours of active high frequency stimulation), and the strength of baseline connectivity. Solid lines correspond to linear model fit (dashed lines mark the 95% confidence intervals) for each individual (Patient 1 had bilateral thalamic stimulation and left and right hemispheres treated independently). Plot shows recording electrodes with statistically significant baseline evoked potentials (paired T-test comparing amplitude [20, 300] ms post-single pulse stimulus to [-480, - 200] ms pre-single pulse stimulus. Voltage traces undergo baseline correction by subtracting the median value for window [-200, -50] ms pre stimulus, and so this baseline period was omitted)). BSEP=brain stimulation evoked potentials. HF-DBS = high frequency deep brain stimulation. AM=anteromedial; AV=anteroventral; AD=anterodorsal; MD=mediodorsal; VApc=ventral anterior, parvocellular division; VLa=ventrolateral anterior nucleus; VLpd=ventrolateral posterior nucleus, dorsal part; VLpv=ventrolateral posterior nucleus, ventral part; LD=laterodorsal; CL=central lateral nucleus PuM=medial pulvinar; PuL=lateral pulvinar.

### Thalamocortical effective connectivity is suppressed after >1.5 hours of high frequency DBS

Patients underwent HF-DBS for varying durations, and the active phase of duty-cycle HF-DBS was computed for each. For example, Patient 4, with a 1-minute on / 3-minute off duty-cycle received 4.3 hours of active HF-DBS with 12.9 hours of off-phase time. Representative evoked potentials at baseline and post HF-DBS are shown for two subjects (Fig. 2B). For Patient 2, who received less than 1.5 hours of stimulation, the evoked potential amplitude remained similar between baseline and post HF-DBS. However, for Patient 3, who received >1.5 hours of HF-DBS, there was a clear reduction in evoked potential amplitude.

Figure 2C shows a clear separation in induced changes in BSEP amplitude based on the HF-DBS duration. Stimulation lasting more than 1.5 hours consistently resulted in suppression of BSEP amplitude (negative Cohen’s *d*), while shorter durations showed no effect or enhancement (zero or positive Cohen’s d). Furthermore, the effects of modulation were greatest for areas with stronger baseline connectivity (Fig. 2C).

A linear regression model comparing baseline BSEP amplitudes and the degree of HF-DBS induced modulation revealed different responses between short- and long-duration HF-DBS groups (P = 0.0016; 95% confidence interval of the difference in mean slope = [-1.53, -0.48]).

Thus, baseline effective connectivity predicted the effect size of BSEP suppression following >1.5 hours of HF-DBS. Patient-specific regression model metrics are in Supplemental Table 3.

### The effects of thalamic high frequency DBS are network specific

The anatomical distributions of thalamocortical BSEPs are shown in Figure 3. The distribution of BSEPs generally overlapped with expected neuroanatomical networks, with greater BSEP amplitudes seen in contacts within vs. outside of expected regions (P<1x10^-10^) (Fig. 3B/C).

Supplemental Figures 1 – 3 show electrode position relative to labeled nuclei in multiple planes, modeled tissue activation volumes, and BSEPs amplitudes projected onto electrode contact positions, respectively. Due to the sample size and the overlap of activated tissue volumes across multiple structures, a highly specific mapping of thalamic subfield connectivity was not possible. However, some general patterns of engagement are evident across thalamic functional groups.

Stimulation of the motor thalamus (ventral group including the ventrolateral (VL) and ventral anterior (VA) nuclei) elicited responses in motor/premotor, and prefrontal (more so with VA) regions, while lacking reliable engagement of limbic, temporal neocortical, and posterior structures (Patients 5, 8, and 9 with ventral group stimulation alone; Patients 1, 2, 3, 6, and 7 with ventral group plus anterior complex stimulation; Patient 10 with anterior complex and mediodorsal nucleus). Medial pulvinar BSEPs (from a single participant, Patient 4) were most evident in posterior quadrant and temporal neocortical regions, with lesser but perceptible impact on some frontal and insula regions; a high burden of IEDs may have increased the variance of background activity and reduced BSEP effect size. Patients 1, 2, 3, 6, 7, and 10 had volumes of activated tissue engaging the limbic thalamus (anterior complex, including the anteroventral (AV) and anteromedial (AM) nuclei or mammillothalamic tract (MTT), and mediodorsal nucleus); all members of this group additionally have motor thalamus engagement (VA nucleus), except Patient 10. This group had particularly high amplitude and widely distributed BSEPs, broadly engaging frontotemporal regions. Anterior complex/VA stimulation elicited statistically significant BSEPs in 257 of 434 ipsilateral cortical contacts (59%), compared to 84 of 512 (16%) in the remaining cohort, while median BSEP amplitude was 1.84 and 0.79 SD units above background activity, respectively (P<1x10^-10^), Supplemental Figure 4, with higher amplitudes seen in frontal vs. temporal regions.

Figure 3D and Supplemental Figure 3B shows the distribution of thalamic HF-DBS-induced changes in effective connectivity, with stimulation-duration dependence (long-duration DBS (>1.5 hours) noted by red and short-duration (<1.5 hours) by blue outlines; listed HF-DBS duration is the total time of the active phase of duty-cycle stimulation). The correlation between baseline connectivity strength and DBS-induced modulation of BSEP amplitudes, shown in Figure 2, is evident in these maps. Patient 6 is an exemplary case of duration-dependent modulation, with suppression of BSEP amplitudes clearly seen after 5.8 hours of HF-DBS, in contrast to weak 1 hour effects (selected BSEP voltage tracings shown in Supplemental Figure 5). It is also notable that beyond a lack of BSEP suppression, there was in fact apparent enhancement of BSEP amplitudes with short-duration stimulation, which was correlated with baseline connectivity strength (Supplemental Table 3).

### Thalamic high frequency DBS modulates interictal epileptiform discharges in a network-selective manner

Figures 2-3 show that thalamocortical BSEPs were modulated by HF-DBS in a manner that was highly network specific and influenced by baseline BSEP strength. To directly assess the relevance of these findings for epilepsy, we assessed the impact of HF-DBS on IEDs—a well-established biomarker of epilepsy network activity—and characterized the association between baseline thalamocortical effective connectivity and IED rate modulation. Interestingly, the effect of HF-DBS on IEDs was immediate and was observable between the on- and off-phases of duty-cycle stimulation for some individuals (Fig. 4A), in contrast to the delayed modulation of BSEP amplitude.

**Figure 3.**
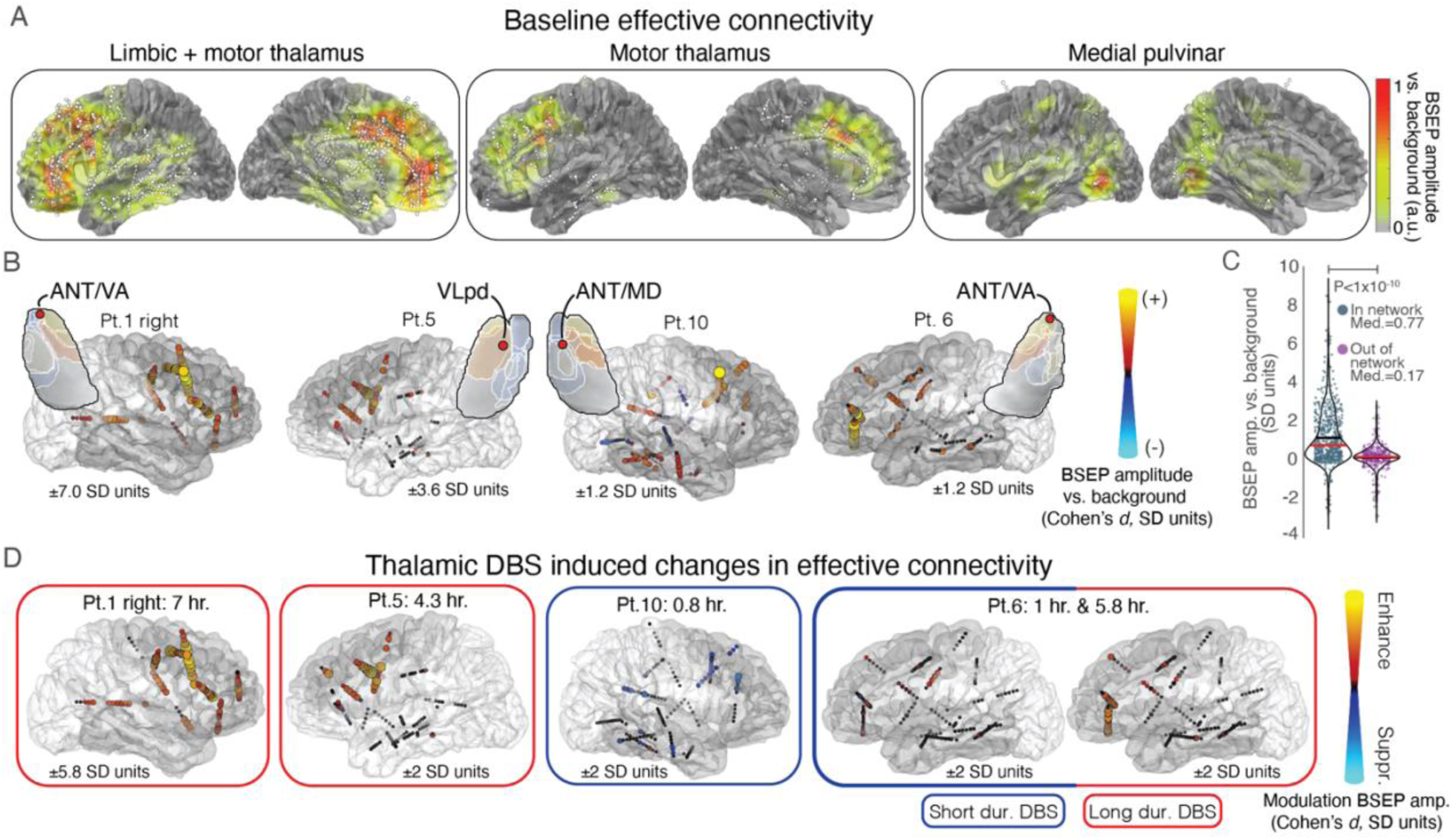
High frequency thalamic stimulation modulates brain excitability, with network specificity and stimulation duration dependence. A) Thalamocortical effective connectivity heatmaps were generated across patients, grouped into limbic + motor thalamus (n=7; Patient 10 with limbic-only stimulation included here), motor thalamus alone (n=3), and medial pulvinar (n=1) categories. B) Representative baseline BSEP amplitude is encoded in a color map and overlayed on ipsilateral electrode contacts. Regions with expected anatomical connectivity^26^ to stimulated thalamic structures are marked in dark grey—brain parcels were clustered into orbitofrontal, prefrontal, cingulate, insula, neocortical temporal, mesial temporal, parietal, occipital, precentral, and postcentral regions. Insets show axial thalamic slices (anatomical orientation), the stimulated cathodal contact (red circles), and relevant nuclei (detailed labeling in Supplementary Figure 1; all subject renderings in Supplementary Figure 3). Contact-to-thalamic structure distance is detailed in Supplemental Fig. 1 – 2 and Table 1. C) Violin plot of BSEP amplitude (Cohen’s *d*) for contacts within versus outside of expected thalamocortical networks. Black and red horizontal lines indicate mean and median values, respectively. D) Representative thalamic HF-DBS induced changes in network effective connectivity (comparing baseline and post HF-DBS BSEP amplitude), corresponding to thalamic targets as shown in B. Consistent suppression of effective connectivity is seen with HF-DBS duration >1.5 hours (red border) vs. <1.5 hours (blue border) (listed durations are the active phase of duty-cycle stimulation). The correlation between baseline connectivity strength and DBS-induced modulation of BSEP amplitude, shown in Figure 2, is evident in these maps. BSEP = brain stimulation evoked potential. HF-DBS = high frequency deep brain stimulation. SD = standard deviation. AM=anteromedial; AV=anteroventral; MTT=mammillothalamic tract; VA= ventral anterior; VL=ventrolateral; PuM=medial pulvinar.

**Figure 4.**
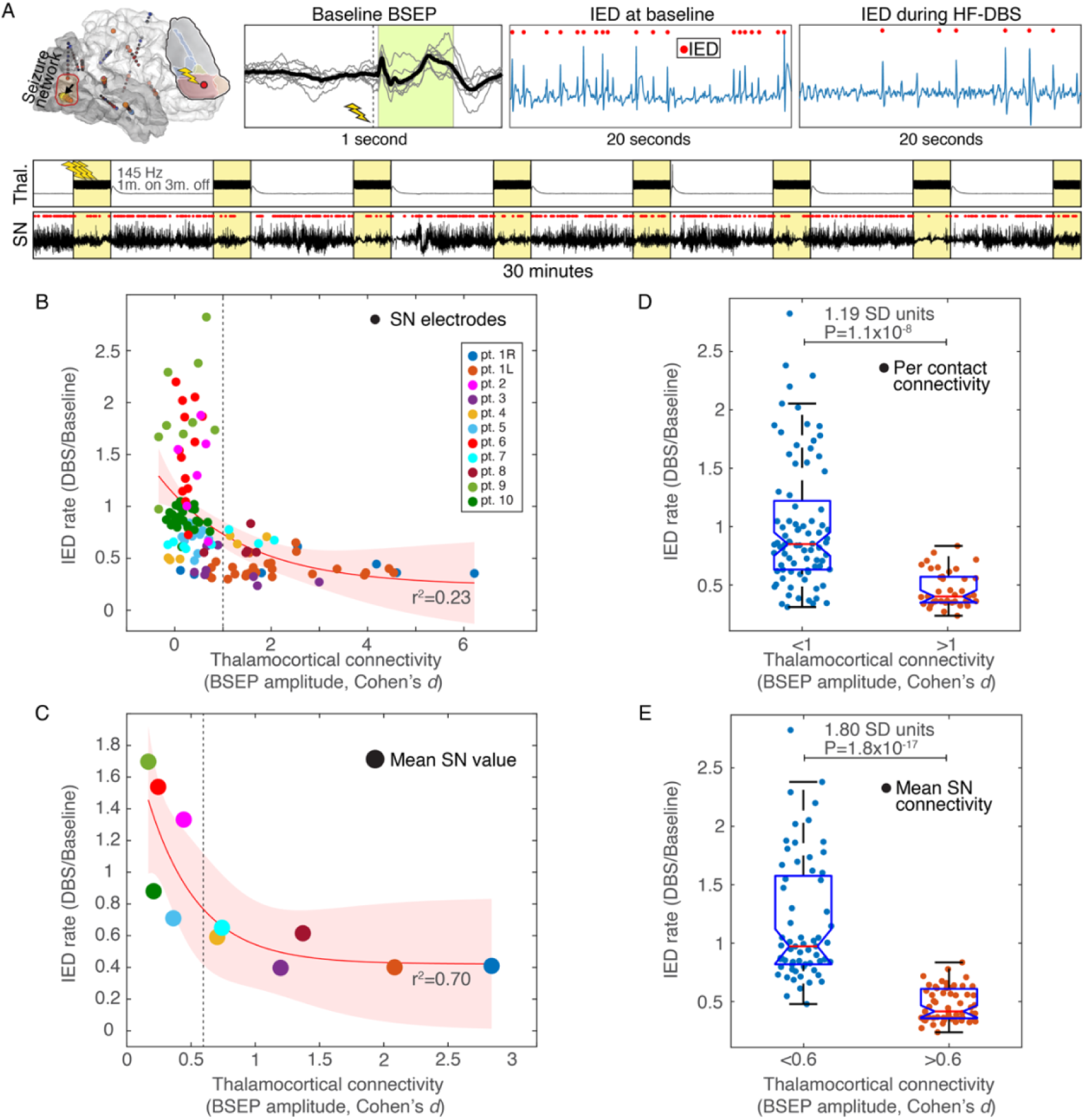
Thalamocortical connectivity predicts the impact of HF-DBS on epileptiform activity (seizure network excitability). A) Baseline BSEP amplitude quantified thalamocortical connectivity. IEDs were quantified using a validated classifier in the awake state, at baseline and during HF-DBS. The lightning bolt marks the pulvinar stimulation site; the black arrow marks the channel corresponding to displayed voltage tracings. The bottom panels of A) show 30 minutes of sEEG data from the thalamus and SN during duty-cycle HF-DBS, with evident modulation of IEDs during each stimulation on-phase; representative data from Pt. 4. B) Scatter plot of thalamic HF-DBS induced change in the rate of IED rate, relative to baseline thalamocortical BSEP amplitude, measured from each SN electrode contact; C) corresponding mean SN values, both with exponential fit model [*y*(*x*) = *a* ∗ *e* ^(−*b∗x*)^ + *c*] with 95% confidence interval and coefficient of determination. Y-axis values are the ratio of the IED rate during HF-DBS relative to baseline (unitless). D) Box-plot of IED rate modulation per-contact, binarized into low and high per-contact connectivity groups, and E) low and high average SN connectivity groups. *Patient 7 results from rapid-cycling DBS (10 sec. on 10 sec. off). Three descriptive models of how thalamocortical connectivity may predict the impact of thalamic HF-DBS on the rate of IEDs are shown in Supplemental Figure 6. BSEP = brain stimulation evoked potential. HF-DBS = high frequency deep brain stimulation; SN = seizure network; IED = interictal epileptiform discharge.

### Response to chronic neuromodulation

Three study participants proceeded to chronic neuromodulation and had > 3-month follow up (Supplementary Table 4). Patients 4 and 7, with favorable BSEP and IED rate modulation, had marked clinical benefit—74% and 100% seizure rate reduction, undergoing 4-lead ANT + Pulvinar DBS, and ANT-DBS (the additional cingulate leads in this 4-lead construct have not been activated), respectively. In contrast, Patient 1 had clear awake-state biomarker suppression, but no reduction in seizure frequency from sequential VNS and ANT-DBS therapy. Post-hoc analysis of the thalamic sEEG HF-DBS trial revealed negligible IED modulation during sleep (Supplementary Figure 7); of note, primary semiology for this subject was nocturnal hypermotor seizures.

## Discussion

Rapid electrophysiological biomarkers are needed to assess network engagement and track excitability for neuromodulation of epilepsy and disorders with diverse networks and delayed clinical effects. Our findings identify such DBS biomarkers in a cohort of individuals with drug-resistant epilepsy undergoing stereotactic EEG, and provide key insights into the short-latency effects of thalamic DBS. First, baseline thalamocortical BSEPs delineate distinct patterns of network engagement between neighboring thalamic subfields. Second, changes in effective connectivity depend on the duration of stimulation, with reliable suppression of network excitability—measured by change in BSEP amplitude—occurring after >1.5 hours of active HF-DBS (>780,000 pulses), consistent with neuroplasticity-mediated effects. Third, thalamic HF-DBS induced changes in network excitability depend on the baseline strength of thalamocortical connections, which can be mapped using BSEPs. Fourth, acute modulation of pathological interictal SN activity (IED rate) by HF-DBS depends on overall SN engagement—this acute modulation of IED rate is consistent with direct electrical effects of stimulation.

The electrophysiological biomarkers characterized in this study may provide a pragmatic approach for individualized optimization of DBS in epilepsy. Trials of thalamic DBS, combined with repeated BSEP measurements and IED quantification can demonstrate SN engagement, quantify acute electrical modulation of pathological SN activity (IEDs), and assess subacute changes in network excitability (e.g. plasticity), for efficient data-driven tuning of DBS^28^.

Importantly, previous work with transcranial magnetic stimulation^29^ and responsive neurostimulation^30^ has shown that stimulation-evoked measures of excitability and IED rate are correlated with long-term seizure control, suggesting these acute/subacute biomarkers may inform response to chronic neuromodulation. Furthermore, this work suggests that electrode targeting to optimize SN engagement may be preferred over a generic “sweet-spot.” However, further prospective work in larger patient cohorts will be needed to determine the clinical predictive value of acute BSEP and IED modulation for long-term seizure control, and to advance best practices to identify DBS targets and stimulation parameters.

Establishing electrophysiological biomarkers of network excitability is particularly timely given the recent FDA approval of a sensing-capable, adaptive DBS system (ClinicalTrials.gov ID NCT04547712, “Adaptive DBS Algorithm for Personalized Therapy in Parkinson’s Disease (ADAPT-PD)”)^31^. While the ADAPT-PD study provides proof of concept of biomarker driven adaptive DBS, additional device capabilities will be needed for epilepsy biomarkers described here, and we hope this work can inform design decisions for next-generation epilepsy neuromodulation systems. Design features would optimally include 4-lead constructs to allow for lead placement in subcortical and cortical SN nodes, single pulse stimulation delivery and recordings to track BSEP-based measures of connectivity and excitability, timeseries recordings and feature extraction to quantify IEDs (and seizures), and program flexibility to screen, quantify, and tune stimulation parameters. Next generation systems would benefit from bidirectional connectivity to an off-body device for data transfer to the cloud and remote programming, such as provided by existing investigational 4-lead systems^32^. This biomarker targeted stimulation would allow for data-driven stimulation contact selection and programming optimization with the goal of seizure control.

The relatively long duration of HF-DBS required to suppress thalamocortical effective connectivity suggests neuroplasticity-based changes in neuronal activity, such as homeostatic plasticity^33,34^. Homeostatic plasticity mechanisms, such as synaptic scaling^35^ and regulation of intrinsic excitability^36^, stabilize network activity and neuron firing rates when exposed to a perturbation that disrupts the existing equilibrium. These mechanisms operate over hours- to days-long timescales^37,38^, consistent with the timescale of DBS-induced modulation observed here, and parallel the clinical effects of DBS seen in disorders such as Tourette syndrome (motor tics), Parkinson’s disease (bradykinesia), central pain, and mood disorders^6^. Thus, HF-DBS may trigger a homeostatic response that reduces SN excitability which can ultimately lower the seizure burden. This is in contrast to the relative accentuation of BSEP amplitudes seen with HF-DBS durations <1.5 hours. These acute excitatory effects of HF-DBS align with previous findings on HF stimulation of cortical and subcortical white matter structures^39^.

Whereas HF-DBS reliably suppressed network excitability only after about 1.5 hours of stimulation, HF-DBS acutely suppressed SN IED rate. These two electrophysiological biomarkers appear to characterize different aspects of network modulation. The acute effects of HF-DBS on IEDs seen here have been linked to the desynchronization of local field activity^40^, in contrast to subacute change in network excitability mediated by neuroplastic mechanisms.

Similarly, the immediate tremor improvements seen with DBS for essential tremor and Parkinson’s disease have been attributed in part to “informational lesioning” or “jamming” of pathological oscillatory circuit activity^41,42^, in contrast to neuroplasticity-mediated modulation of bradykinesia, dystonia, and motor tics^6^. Biomarkers reflecting distinct mechanisms of network modulation—namely direct electrical network desynchronization assessed by IED rate, and subacute excitability change assessed by BSEPs—may offer complementary insights into the effects of DBS to inform parameter optimization.

The SN IED rate modulation was dependent on overall SN engagement, as measured by baseline thalamocortical BSEP amplitude across SN contacts (Fig. 4, Supplemental Fig. 6). This aligns with a network model of epilepsy^7–10^ in which sufficient engagement of network nodes can modulate the broader pathological network^8^, including subregions with poor thalamocortical connectivity. Additionally, these results provide unique electrophysiological evidence supporting a previously proposed “network theory” of ANT-DBS^43^, consistent with the connectivity dependence of DBS efficacy for other neurological disorders^44–47^. This study underscores the need for continued investigation into network-targeted neuromodulation strategies for epilepsy^10^. Moreover, biomarkers of network engagement and modulation may hold translational potential for other disorders of aberrant network excitability with delayed clinical response, including neuropsychiatric conditions, dystonia, central pain, and stroke recovery.

This study also provides strong evidence that the anatomical distribution of DBS effects differs between neighboring thalamic nuclei (Fig. 3). It is important to note that these stimulation derived connectivity maps provide information that is distinct from and complementary to thalamic seizure recordings^48,49^. This is a critical point—corticothalamic and thalamocortical effective connectivity is not necessarily reciprocal^50^, and it may follow that the thalamic nucleus with the earliest or maximal ictal activity is not the DBS target that best engages the SN.

Relatively higher amplitude and more broadly distributed BSEPs were observed when stimulation fields engaged both the anterior complex (AM nucleus, AV nucleus, and MTT) and the VA nucleus, engaging limbic frontotemporal, and motor and diffuse prefrontal regions, respectively. This finding provides an effective connectomics rationale for the network-theory of ANT-DBS^43^, and complements prior empirical “sweet-spot” analyses of ANT-DBS^51–53^. It is unclear if the higher amplitude and more broadly distributed BSEPs elicited from the anterior complex/VA region results directly from stimulated nuclei, engagement of the anterior thalamic radiations, or alternative and patient specific factors. Ultimately, selecting a DBS target that maximizes engagement of an individual’s SN may yield better outcomes than using generic stimulation targets.

The posterior quadrant and temporal neocortical predominance of BSEPs elicited by medial pulvinar stimulation is drawn from a single patient, and more work is needed to determine the representativeness of results. These findings can, however, be interpreted in the context of existing work from our group^54^ and others^50^. The predominance of neocortical temporal over mesial temporal engagement by pulvinar BSEPs is consistent with prior work, which has demonstrated that connectivity is not always bidirectional^50^ (strong mesial temporal to medial pulvinar effective connectivity but sparse medial pulvinar to mesial temporal connectivity; strong bidirectional medial pulvinar and temporal neocortical connectivity). These findings suggest that pulvinar BSEPs can identify patterns of connectivity that are complementary to passive sEEG recordings^55^. It is also important to note that subregions within the medial and lateral pulvinar have distinct connectivity profiles^56,57^, which are relevant for individualized pulvinar DBS targeting. Connectivity mapping from Patient 4’s ventromedial pulvinar lead does not contradict prior work showing frontal, parietal, and cingulate coupling to the dorsomedial pulvinar, or early visual and extrastriate occipital cortex coupling to the lateral pulvinar^54,56,57^. The single patient with pulvinar stimulation therefore serves as an example case of how the proposed biomarkers may be used beyond ANT stimulation.

This study is limited by the relative rarity of human thalamic sEEG and the complexity of clinical care in the epilepsy monitoring unit. The HF-DBS trials were delivered as a part of clinical care, contributing to variability in thalamic targets, stimulation parameters, and HF-DBS duration. One patient had resumption of antiseizure medications between the baseline and post-DBS BSEP measurements, which may have impacted brain excitability. Stimulation was delivered using a voltage clamped external neurostimulator, and all patients received between 4- 6 V stimulation amplitude (except Patient 2 who had a short stimulation window and underwent continuous DBS at 7V in an attempt to deliver a clinically impactful stimulation trial); differences in electrode impedances contributed to the spread of delivered currents. While there was inter-subject variability in stimulation amplitude and duty-cycle, we did not identify a consistent association between these settings differences and BSEP or IED modulation.

While this acute sEEG biomarker study establishes that BSEPs and short DBS trials capture effective connectivity and immediate network consequences of thalamic stimulation, prospective clinical trials are now required to confirm the predictive value of such biomarkers for long-term seizure reduction, characterize the wash-in and wash-out dynamics of DBS and SN excitability over multiple timescales, and to refine best practices for biomarker informed neuromodulation^31^ (chronic DBS outcomes limited by small the sample size (n=3) and larger prospective trials are needed). Future prospective studies should aim for stable stimulation parameter settings to limit potential confounds, and we suggest stimulation amplitudes of about 4-5 mA (as tolerated) and 1 min. on 3 min. off duty cycle (balancing efficiency of stimulation delivery, tolerability, and typical clinical practice habits^58^). Ideally, we envision a two-stage workflow: 1) sEEG phase with BSEP and short DBS trials to screen and identify favorable stimulation targets and parameters to engage and suppress the SNs, and 2) ambulatory phase, with periodic BSEP/IED assessments to tune settings over time (this will require next-generation sensing IPGs with expanded capabilities, as outlined above). Importantly, because this strategy hinges on generic principles of network engagement and excitability, this could be extended to a range of neurologic and psychiatric brain circuit disorders where delayed or subjective clinical outcomes makes iterative optimization difficult. Stage 1) alone would allow for individualization of stimulation target selection and parameter optimization, until next-generation IPGs become available.

This paper limited analysis of BSEPs and IEDs to the awake state. This was done to avoid known wake/sleep state dependent changes in IED rate^12^ and effective connectivity^59^. Given the discordance in wake/sleep state IED modulation in Patient 1, future work may benefit from consideration of behavioral state dependent effects. The lack of response to chronic VNS and ANT-DBS for Patient 1 may also suggest a particularly challenging epilepsy phenotype in this patient.

The spatial under-sampling of sEEG is a fundamental limitation of thalamic sEEG, and precludes definitive identification of the ‘ideal’ nucleus of every SN (note poor SN engagement in patients 2, 5, 6, 9, 10, Fig. 4C). Pre-sEEG clinical assessment combined with existing knowledge of thalamus connectivity should guide the selection of putative thalamic seizure network nodes for targeting. Also of note, we analyzed BSEP amplitude over 20 – 300 milliseconds post-stimulus to avoid stimulation artifact and to encompass the canonical N1/N2 response range, which is believed to include oligo- and polysynaptic propagation^13^. While this BSEP window is not limited to monosynaptic connections, the presence of a reliable BSEP does indicate effective connectivity—the stimulated thalamic site exerts an influence on the recorded region. This is important when interpreting the spatial extent of engagement, nevertheless, the modulation of BSEP amplitude after HF-DBS supports its use as a quantitative marker of network excitability.

Here, we combined single pulse and repetitive HF thalamic stimulation during sEEG to elucidate the effects of thalamic stimulation on brain network dynamics across multiple timescales. Our findings underscore the central role of SN engagement for DBS induced modulation of IEDs and SN excitability. The practical, accessible electrophysiological biomarkers identified here may support a new paradigm of biomarker informed DBS, with SN-specific electrode targeting, and data-driven tuning of stimulation parameters, to improve patient care.

## Data Availability

All data produced in the present study are available upon reasonable request to the authors

## Acknowledgement

This work was supported by the National Institute of Neurological Disorders and Stroke awards K23NS136792, R01NS09288203, and National Institute of Mental Health award R01MH122258—the content is solely the responsibility of the authors and does not represent the official views of the NIH—the Tianqiao and Chrissy Chen Career Development Award in Translational Research (Neuro), and the CLARA project that has received funding from the European Union’s HORIZON EUROPE research and innovation program, Grant Agreement No 101136607.

## Author Contribution**s**

N.M.G., D.H., G.A.W contributed to the conception and design of the study. N.M.G., D.H., G.A.W., T.P, G.O.V, H.H., K.J.M., B.N.L, J.J.VG, and V.K. contributed to the acquisition and analysis of data. N.M.G, D.H., and T.P. contributed to drafting the text or preparing the figures.

## Data availability

Data and code to reproduce the main findings of this study are available on GitHub (https://github.com/nmgregg/Thalamic-stim-connect). Specific inquiries can be sent to the corresponding author.

## Supplementary Materials

Supplementary material is available online.

## References

1. Schuepbach WM, Rau J, Knudsen K, et al. Neurostimulation for Parkinson’s disease with early motor complications. N Engl J Med. 2013;368(7):610–622.

2. Fisher R, Salanova V, Witt T, et al. Electrical stimulation of the anterior nucleus of thalamus for treatment of refractory epilepsy. Epilepsia. 2010;51(5):899–908.

3. Boccard SG, Pereira EA, Aziz TZ. Deep brain stimulation for chronic pain. J Clin Neurosci. 2015;22(10):1537–1543.

4. Tisch S, Rothwell JC, Bhatia KP, et al. Pallidal stimulation modifies after-effects of paired associative stimulation on motor cortex excitability in primary generalised dystonia. Exp Neurol. 2007;206(1):80–85.

5. Alagapan S, Choi KS, Heisig S, et al. Cingulate dynamics track depression recovery with deep brain stimulation. Nature. 2023;622(7981):130–138.

6. Ashkan K, Rogers P, Bergman H, Ughratdar I. Insights into the mechanisms of deep brain stimulation. Nat Rev Neurol. 2017;13(9):548–554.

7. Morgan RJ, Soltesz I. Nonrandom connectivity of the epileptic dentate gyrus predicts a major role for neuronal hubs in seizures. Proc Natl Acad Sci U S A. 2008;105(16):6179–6184.

8. Wilke C, Worrell G, He B. Graph analysis of epileptogenic networks in human partial epilepsy. Epilepsia. 2011;52(1):84–93.

9. Richardson MP. Large scale brain models of epilepsy: dynamics meets connectomics. J Neurol Neurosurg Psychiatry. 2012;83(12):1238–1248.

10. Piper RJ, Richardson RM, Worrell G, et al. Towards network-guided neuromodulation for epilepsy. Brain. 2022;145(10):3347–3362.

11. Fisher RS, Acevedo C, Arzimanoglou A, et al. ILAE official report: a practical clinical definition of epilepsy. Epilepsia. 2014;55(4):475–482.

12. Karoly PJ, Rao VR, Gregg NM, et al. Cycles in epilepsy. Nature Reviews Neurology. 2021.

13. Keller CJ, Honey CJ, Megevand P, Entz L, Ulbert I, Mehta AD. Mapping human brain networks with cortico-cortical evoked potentials. Philos Trans R Soc Lond B Biol Sci. 2014;369(1653).

14. van Blooijs D, van den Boom MA, van der Aar JF, et al. Developmental trajectory of transmission speed in the human brain. Nat Neurosci. 2023;26(4):537–541.

15. Keller CJ, Huang Y, Herrero JL, et al. Induction and Quantification of Excitability Changes in Human Cortical Networks. J Neurosci. 2018;38(23):5384–5398.

16. Valentin A, Alarcon G, Honavar M, et al. Single pulse electrical stimulation for identification of structural abnormalities and prediction of seizure outcome after epilepsy surgery: a prospective study. Lancet Neurol. 2005;4(11):718–726.

17. Matsumoto R, Kunieda T, Nair D. Single pulse electrical stimulation to probe functional and pathological connectivity in epilepsy. Seizure. 2017;44:27–36.

18. Huang H, Ojeda Valencia G, Gregg NM, et al. CARLA: Adjusted common average referencing for cortico-cortical evoked potential data. J Neurosci Methods. 2024;407:110153.

19. Janca R, Jezdik P, Cmejla R, et al. Detection of interictal epileptiform discharges using signal envelope distribution modelling: application to epileptic and non-epileptic intracranial recordings. Brain Topogr. 2015;28(1):172–183.

20. Sladky V, Nejedly P, Mivalt F, et al. Distributed brain co-processor for tracking spikes, seizures and behaviour during electrical brain stimulation. Brain Commun. 2022;4(3):fcac115.

21. Dell KL, Payne DE, Kremen V, et al. Seizure likelihood varies with day-to-day variations in sleep duration in patients with refractory focal epilepsy: A longitudinal electroencephalography investigation. EClinicalMedicine. 2021:100934.

22. Horn A, Li N, Dembek TA, et al. Lead-DBS v2: Towards a comprehensive pipeline for deep brain stimulation imaging. Neuroimage. 2019;184:293–316.

23. Krauth A, Blanc R, Poveda A, Jeanmonod D, Morel A, Szekely G. A mean three-dimensional atlas of the human thalamus: generation from multiple histological data. Neuroimage. 2010;49(3):2053–2062.

24. Fischl B. FreeSurfer. Neuroimage. 2012;62(2):774–781.

25. Destrieux C, Fischl B, Dale A, Halgren E. Automatic parcellation of human cortical gyri and sulci using standard anatomical nomenclature. Neuroimage. 2010;53(1):1–15.

26. Jones EG. The Thalamus. Second ed. Cambridge: Cambridge University Press; 2007.

27. Amunts K, Lepage C, Borgeat L, et al. BigBrain: an ultrahigh-resolution 3D human brain model. Science. 2013;340(6139):1472–1475.

28. Stieve BJ, Richner TJ, Krook-Magnuson C, Netoff TI, Krook-Magnuson E. Optimization of closed-loop electrical stimulation enables robust cerebellar-directed seizure control. Brain. 2023;146(1):91–108.

29. Pawley AD, Chowdhury FA, Tangwiriyasakul C, et al. Cortical excitability correlates with seizure control and epilepsy duration in chronic epilepsy. Ann Clin Transl Neurol. 2017;4(2):87–97.

30. Arcot Desai S, Tcheng TK, Morrell MJ. Quantitative electrocorticographic biomarkers of clinical outcomes in mesial temporal lobe epileptic patients treated with the RNS(R) system. Clin Neurophysiol. 2019;130(8):1364–1374.

31. Oehrn CR, Cernera S, Hammer LH, et al. Chronic adaptive deep brain stimulation versus conventional stimulation in Parkinson’s disease: a blinded randomized feasibility trial. Nat Med. 2024.

32. Kremen V, Sladky V, Mivalt F, et al. Modulating limbic circuits in temporal lobe epilepsy: impacts on seizures, memory, mood and sleep. Brain Commun. 2025;7(2):fcaf106.

33. Turrigiano GG. The self-tuning neuron: synaptic scaling of excitatory synapses. Cell. 2008;135(3):422–435.

34. Chai Z, Ma C, Jin X. Homeostatic activity regulation as a mechanism underlying the effect of brain stimulation. Bioelectron Med. 2019;5:16.

35. Turrigiano GG, Nelson SB. Homeostatic plasticity in the developing nervous system. Nat Rev Neurosci. 2004;5(2):97–107.

36. Marder E, Goaillard JM. Variability, compensation and homeostasis in neuron and network function. Nat Rev Neurosci. 2006;7(7):563–574.

37. Ibata K, Sun Q, Turrigiano GG. Rapid synaptic scaling induced by changes in postsynaptic firing. Neuron. 2008;57(6):819–826.

38. Watt AJ, Desai NS. Homeostatic Plasticity and STDP: Keeping a Neuron’s Cool in a Fluctuating World. Front Synaptic Neurosci. 2010;2:5.

39. Mohan UR, Watrous AJ, Miller JF, et al. The effects of direct brain stimulation in humans depend on frequency, amplitude, and white-matter proximity. Brain Stimul. 2020;13(5):1183–1195.

40. Yu T, Wang X, Li Y, et al. High-frequency stimulation of anterior nucleus of thalamus desynchronizes epileptic network in humans. Brain. 2018;141(9):2631–2643.

41. Grill WM, Snyder AN, Miocinovic S. Deep brain stimulation creates an informational lesion of the stimulated nucleus. Neuroreport. 2004;15(7):1137–1140.

42. Lozano AM, Lipsman N, Bergman H, et al. Deep brain stimulation: current challenges and future directions. Nat Rev Neurol. 2019;15(3):148–160.

43. Salanova V, Sperling MR, Gross RE, et al. The SANTE study at 10 years of follow-up: Effectiveness, safety, and sudden unexpected death in epilepsy. Epilepsia. 2021;62(6):1306–1317.

44. Horn A, Reich M, Vorwerk J, et al. Connectivity Predicts deep brain stimulation outcome in Parkinson disease. Ann Neurol. 2017;82(1):67–78.

45. Johnson KA, Duffley G, Anderson DN, et al. Structural connectivity predicts clinical outcomes of deep brain stimulation for Tourette syndrome. Brain. 2020;143(8):2607–2623.

46. Middlebrooks EH, Okromelidze L, Wong JK, et al. Connectivity correlates to predict essential tremor deep brain stimulation outcome: Evidence for a common treatment pathway. Neuroimage Clin. 2021;32:102846.

47. Hollunder B, Ostrem JL, Sahin IA, et al. Mapping dysfunctional circuits in the frontal cortex using deep brain stimulation. Nat Neurosci. 2024;27(3):573–586.

48. Wu TQ, Kaboodvand N, McGinn RJ, et al. Multisite thalamic recordings to characterize seizure propagation in the human brain. Brain. 2023;146(7):2792–2802.

49. Guye M, Regis J, Tamura M, et al. The role of corticothalamic coupling in human temporal lobe epilepsy. Brain. 2006;129(Pt 7):1917–1928.

50. Rosenberg DS, Mauguiere F, Catenoix H, Faillenot I, Magnin M. Reciprocal thalamocortical connectivity of the medial pulvinar: a depth stimulation and evoked potential study in human brain. Cereb Cortex. 2009;19(6):1462–1473.

51. Gross RE, Fisher RS, Sperling MR, Giftakis JE, Stypulkowski PH. Analysis of Deep Brain Stimulation Lead Targeting in the Stimulation of Anterior Nucleus of the Thalamus for Epilepsy Clinical Trial. Neurosurgery. 2021;89(3):406–412.

52. Lehtimaki K, Mottonen T, Jarventausta K, et al. Outcome based definition of the anterior thalamic deep brain stimulation target in refractory epilepsy. Brain Stimul. 2016;9(2):268–275.

53. Schaper F, Plantinga BR, Colon AJ, et al. Deep Brain Stimulation in Epilepsy: A Role for Modulation of the Mammillothalamic Tract in Seizure Control? Neurosurgery. 2020;87(3):602–610.

54. Bilderbeek JA, Gregg NM, Yanez-Ramos MG, et al. Human pulvinar stimulation engages select cortical pathways in epilepsy. bioRxiv. 2025.

55. McGinn R, Von Stein EL, Datta A, et al. Ictal Involvement of the Pulvinar and the Anterior Nucleus of the Thalamus in Patients With Refractory Epilepsy. Neurology. 2024;103(11):e210039.

56. Arcaro MJ, Pinsk MA, Chen J, Kastner S. Organizing principles of pulvino-cortical functional coupling in humans. Nat Commun. 2018;9(1):5382.

57. Arcaro MJ, Pinsk MA, Kastner S. The Anatomical and Functional Organization of the Human Visual Pulvinar. J Neurosci. 2015;35(27):9848–9871.

58. Fasano A, Eliashiv D, Herman ST, et al. Experience and consensus on stimulation of the anterior nucleus of thalamus for epilepsy. Epilepsia. 2021;62(12):2883–2898.

59. Usami K, Korzeniewska A, Matsumoto R, et al. The neural tides of sleep and consciousness revealed by single-pulse electrical brain stimulation. Sleep. 2019;42(6).

