## Supplemental for "Thalamic stimulation induced changes in network connectivity and excitability in epilepsy"

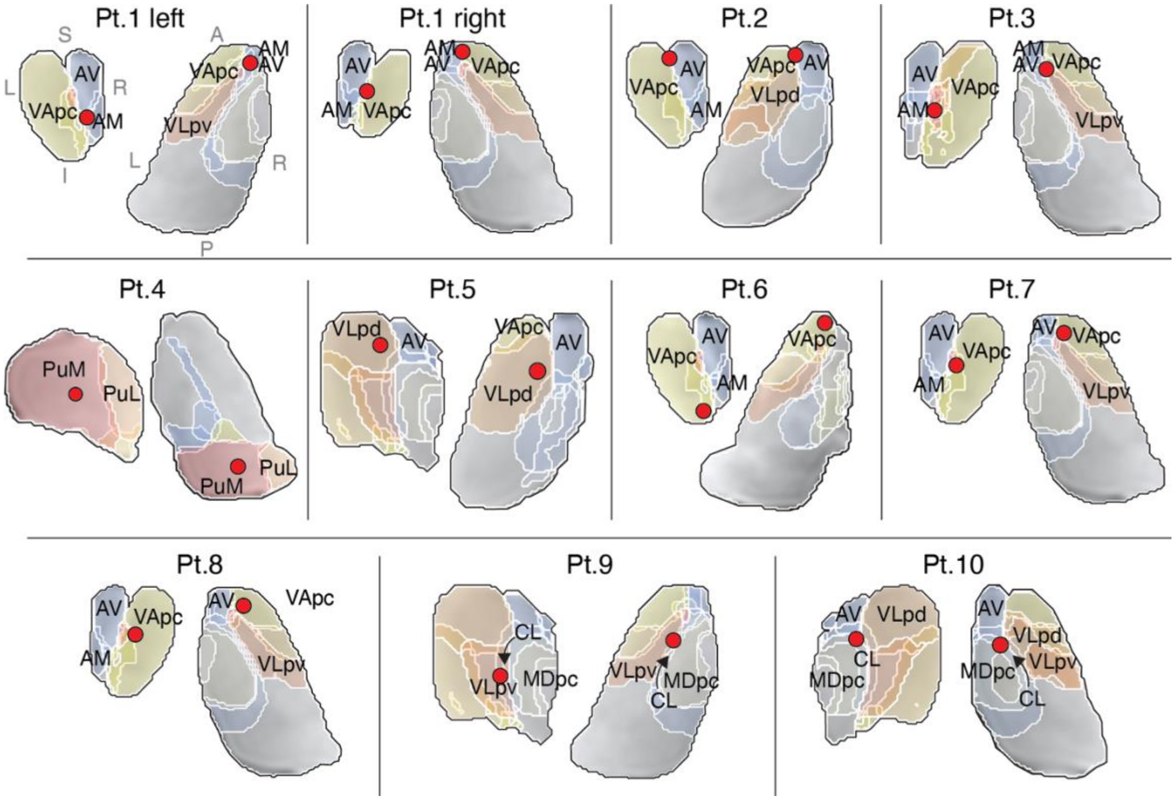


**Supplemental Figure 1. Thalamic electrode localization.** Stimulated cathodal contacts (red circle) and labeled relevant thalamic nuclei in coronal and axial slices from the Krauth/Morel atlas. AM=anteromedial; AV=anteroventral; MTT=mammillothalamic tract; MDpc=mediodorsal, parvocellular part; VAmc=ventral anterior, magnocellular division; VApc=ventral anterior, parvocellular division; VLpd=ventrolateral posterior nucleus, dorsal part; VLpv=ventrolateral posterior nucleus, ventral part; PuM=medial pulvinar; PuL=lateral pulvinar; CL=central lateral nucleus.

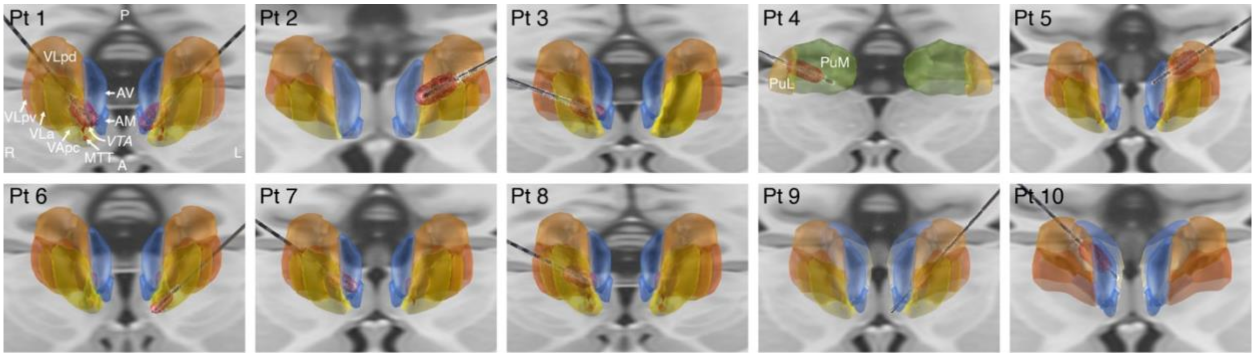


**Supplemental Figure 2. Thalamic stimulation volume of tissue activated (VTA) modeling**. Bipolar VTA models were made using lead DBS v3.2, FieldTrip-SimBio pipeline, (https://www.mrt.uni-jena.de/simbio/index.php/; http://fieldtriptoolbox.org). AM=anteromedial; AV=anteroventral; MTT=mammillothalamic tract; VApc=ventral anterior, parvocellular division; Ventrolateral anterior nucleus; VLpd=ventrolateral posterior nucleus, dorsal part; VLpv=ventral lateral posterior nucleus, ventral part; PuM=medial pulvinar. PuL=lateral pulvinar. *VTA*=volume of tissue activated.

| | Pt. 1 right | Pt. 1 left | Pt 2 | Pt 3 | Pt 4 | Pt 5 | Pt 6 | Pt 7 | Pt 8 | Pt 9 | Pt 10 |
|---|---|---|---|---|---|---|---|---|---|---|---|
| AM | **0.96** | **0.47** | 1.81 | 2.45 | 27.75 | 10.77 | 4.13 | 2.52 | 3.61 | 7.20 | 7.17 |
| AV | **0.28** | **0.33** | **0.31** | **0.95** | 20.71 | 1.96 | 3.94 | **0.89** | 1.85 | 4.52 | **0.86** |
| MTT | 2.17 | 1.72 | 1.72 | **0.24** | 21.86 | 8.60 | **0.81** | **0.38** | 1.30 | 3.69 | 5.49 |
| MDpc | 4.77 | 1.87 | 4.66 | 2.56 | 10.34 | 4.62 | 4.04 | 4.60 | 5.54 | 1.03 | **0.22** |
| VAmc | **0.67** | **0.75** | **0.93** | **0.42** | 21.46 | 8.53 | **0.12** | **0.26** | **0.61** | 4.18 | 6.43 |
| VApc | **0.28** | 1.04 | **0.75** | 1.09 | 20.05 | 4.28 | **0.12** | **0.26** | **0.19** | 2.56 | 4.98 |
| VLpd | 3.37 | 3.25 | 3.08 | **0.91** | 13.57 | **0.11** | 2.18 | 1.37 | 1.71 | 2.24 | 1.36 |
| VLpv | 3.19 | 2.76 | 2.76 | **0.47** | 9.81 | 5.03 | 1.91 | 1.34 | 1.71 | **0.27** | 2.22 |
| PuM | 21.74 | 20.99 | 21.22 | 18.85 | **0.15** | 12.50 | 19.21 | 19.73 | 19.52 | 13.67 | 13.11 |

**Supplemental Table 1. Thalamic electrode localization.** Proximity of stimulated contacts to thalamic nuclei, in millimeters, using the Krauth/Morel atlas. Contacts within 1 mm of a structure are bolded. AM=anteromedial; AV=anteroventral; MTT=mammillothalamic tract; MDpc=mediodorsal parvocellular division; VAmc=ventral anterior magnocellular division; VApc=ventral anterior parvocellular division; VLpd=ventrolateral posterior nucleus, dorsal part; VLpv=ventral lateral ventral part; PuM=medial pulvinar.

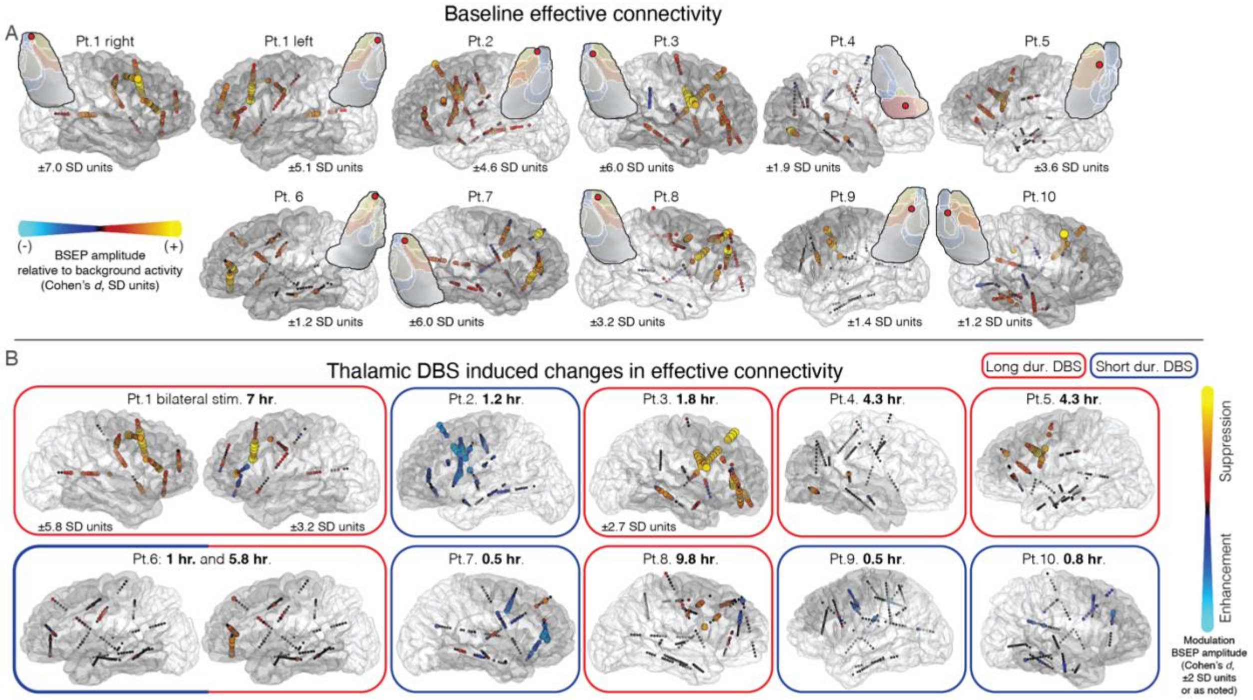


**Supplemental Figure 3. Thalamocortical effective connectivity and high frequency thalamic stimulation induced modulation of network excitability.** A) Baseline brain stimulation evoked potentials (BSEP) with thalamic stimulation and measuring cortical evoked potential amplitude intensity encoded and overlayed on ipsilateral electrode contacts. Cortical parcels and subcortical segments with expected connectivity to stimulated thalamic structures are marked in dark grey, as in main Figure 3. B) Thalamic high frequency stimulation induced changes in network effective connectivity, as in main Figure 3, with consistent suppression of effective connectivity seen with stimulation durations >1.5 hours (red border) vs. <1.5 hours (blue border).

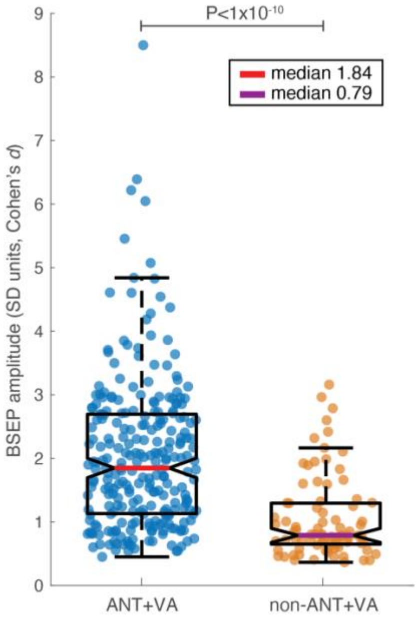


**Supplemental Figure 4. Combined anterior complex and VA nucleus stimulation elicits widely distributed and high amplitude BSEPs.** Boxplots showing brain stimulation evoked potentials (BSEP) amplitude for all ipsilateral cortical contacts with significant evoked responses relative to background activity. ANT+VA group includes patients 1, 2, 3, 6, and 7; non-ANT+VA patients 4, 5, 8, 9, and 10. ANT/VA stimulation elicited statistically significant BSEPs in 257 of 434 ipsilateral cortical contacts (59%), compared to 84 of 512 (16%) in the remaining cohort. Here, ANT referrers to anteromedial and anteroventral nuclei, as well as mammillothalamic tract. ANT=anterior nucleus of the thalamus. VA=ventral anterior nucleus.

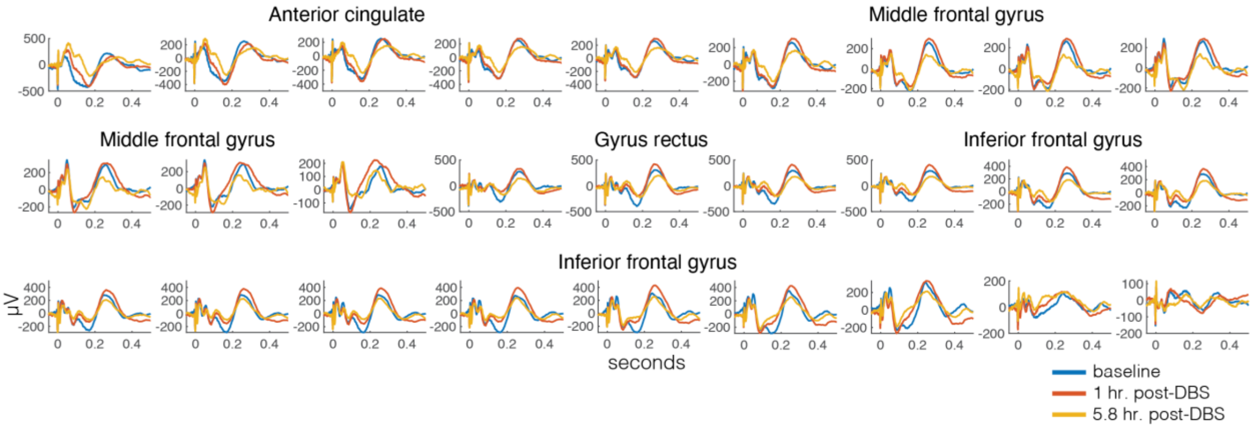


**Supplemental Figure 5. Pulse evoked potentials from each electrode contact along two stereotactic EEG leads.** Pulse evoked potentials at baseline, after 1 hour of high frequency DBS, and after 5.8 hours of high frequency DBS. Data from patient 6.

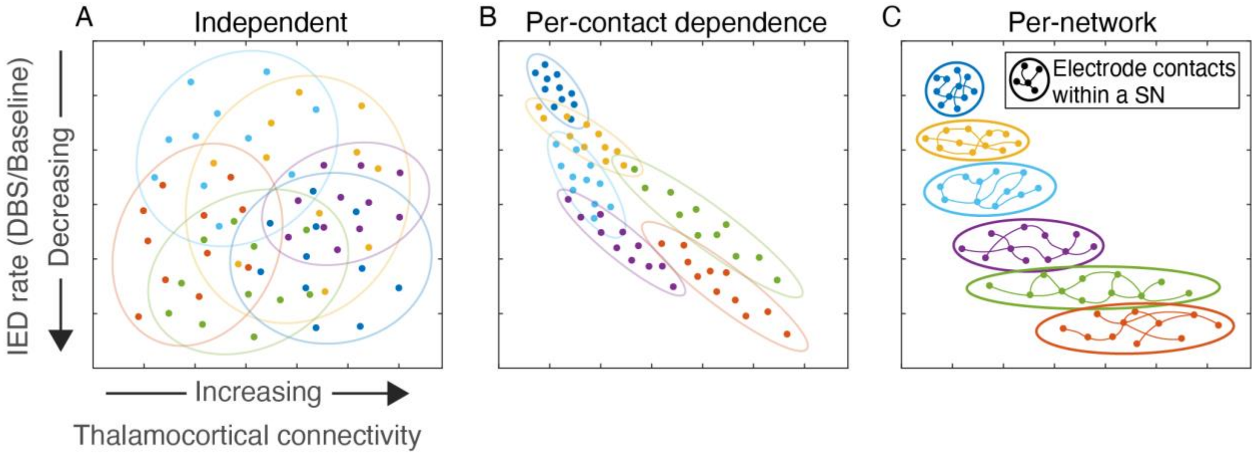


**Supplemental Figure 6. Models of the relationship between thalamocortical connectivity and modulation of IED rate by high-frequency DBS.** A) baseline connectivity amplitude does not predict DBS modulation of IED rate; B) connectivity amplitude is directly predictive of IED rate modulation, per-contact; C) overall SN engagement (average connectivity amplitude across all SN contacts) is predictive of DBS modulation of IED rate. IED = interictal epileptiform discharges. SN = seizure network.

| Patient | Cortical stimulation location | Stimulation parameters |
|---|---|---|
| 6 | Middle & superior temporal gyri | 2 Hz, 200 µs, 1.25 V,<br>384 ohms group impedance |
| 8 | Superior temporal gyrus | 2 Hz, 200 µs, 1.5 V,<br>197 ohms group impedance |

**Supplemental Table 2.** Two patients received cortical low-frequency stimulation that was concurrent with thalamic high frequency DBS, listed in Table 1. Stimulated cortical channels were excluded from pulse evoked potential and interictal epileptiform discharge analyses.

| Patient | DBS duration | Slope | $R^2$ | P-val. of slope coefficient |
|---|---|---|---|---|
| 1 left | **long** | **-0.72** | **0.46** | **1.3E-13** |
| 1 right | **long** | **-0.61** | **0.62** | **3.7E-23** |
| 2 | short | 0.36 | 0.44 | 6.8E-19 |
| 3 | **long** | **-0.41** | **0.35** | **2.5E-12** |
| 4 | **long** | **-0.80** | **0.25** | **4.9E-02** |
| 5 | **long** | **-0.85** | **0.69** | **3.7E-14** |
| 6* | short | 0.46 | 0.18 | 3.2E-03 |
| 6* | **long** | **-1.17** | **0.54** | **7.4E-09** |
| 7 | short | 0.07 | 0.04 | 7.5E-02 |
| 8 | **long** | **-0.15** | **0.04** | **1.5E-01** |
| 9 | short | 1.06 | 0.51 | 2.0E-04 |
| 10 | short | -0.27 | 0.03 | 3.0E-01 |
| **long (median)** | | **-0.72** | **0.46** | |
| short (median) | | 0.36 | 0.18 | |

**Supplemental Table 3.** Linear regression model comparing baseline connectivity strength (pulse evoked potential amplitude) and high-frequency deep brain stimulation (HF-DBS) induced modulation of pulse evoked potential amplitude. Patients were binarized into HF-DBS duration categories: <1.5 hours (short) or > 1.5 hours (long). Linear regression model slope coefficient, $R^2$ goodness of fit, and P-value of the slope coefficient are listed per subject, and median. This table corresponds to manuscript Figure 2. * Patient 6 had pulse evoked potential measurements taken after short and long duration DBS.

| Patient | Intervention details | Follow up duration (mo.) | Surgical epilepsy conference recommendations; interventions; outcomes |
|---|---|---|---|
| 1 | VNS; Bilateral ANT-DBS (Medtronic Percept PC) | 34 (VNS)<br>13 (DBS) | **0% seizure rate reduction.** VNS (20 Hz, 250µs, 2mA, 38% duty-cycle); ANT-DBS (145Hz, 90µs, 7mA total, 1min. on 3min. off). |
| 4 | Bilateral ANT and Pulvinar DBS (Boston Scientific R32) | 20 | **74% seizure rate reduction**. ANT + Pulvinar (143 Hz, 90 µs, 6mA, 1min. on 5min. off to ANT; 7.5mA to pulvinar). Overnight Spike Wave Index 35% baseline → 9% with DBS. |
| 7 | Bilateral ANT and anterior cingulate DBS (Boston Scientific R32) | 18 | **100% seizure rate reduction** (>3 mo. seizure free at last follow up). ANT-DBS (143Hz, 90µs, 5mA, 1min. on 5min. off). Cingulate leads remain off. |

**Supplemental Table 4.** Chronic neuromodulation outcomes for the three subjects that went on to chronic DBS with an implantable device. Electrical current refers to current delivered per-lead. VNS=vagus nerve stimulation. DBS=deep brain stimulation. ANT=anterior nucleus of thalamus.

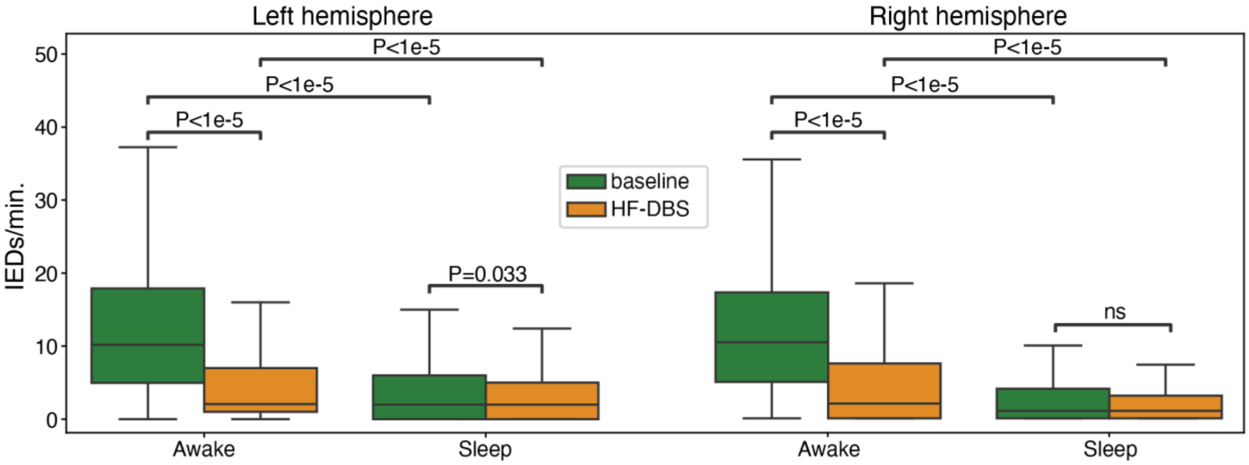


**Supplementary Figure 7.** Patient 1 IED rate in awake and sleep states. IED=interictal epileptiform discharge.
